# Mapping Structural Barriers: A Geospatial Assessment of COVID-19 Vaccine Inequities in Kenya

**DOI:** 10.1101/2025.04.14.25325797

**Authors:** Larry Niño, Agnes Kiragga, F. DeWolf Miller, Samuel M. Mwalili, Godfrey Musuka, Diego F. Cuadros

**Author notes:** Reprints or correspondence: Diego F. Cuadros, Ph.D., Digital Epidemiology Laboratory, University of Cincinnati, Cincinnati, OH, 45221.

## Abstract

**Background:** The COVID-19 pandemic has exacerbated global health inequities, with low-income countries lagging behind in vaccine coverage. By late 2023, only 36% of Kenyans had received at least one vaccine dose, far below the African Union’s 60% target. This study examines spatial disparities in COVID-19 vaccination rates across Kenya, exploring how socioeconomic, environmental, and healthcare infrastructure factors shape vaccine access. Unlike previous studies focused on individual determinants, this research employs a spatial epidemiological approach to uncover structural barriers to equitable vaccination.

**Methods:** This study uses data from the 2022 Kenya Demographic and Health Survey (KDHS), integrating socioeconomic, health, and environmental variables across 1,692 georeferenced clusters. Analytical methods include spatial clustering (K-Means), spatial autocorrelation (Moran’s I), Random Forest regression, and the Erreygers Concentration Index (ECI) to quantify vaccine inequities. A Development Index (DI) was constructed to assess how financial access, living conditions, and healthcare systems influence vaccination rates.

**Results:** Our results reveal stark geographic disparities: vaccination rates range from 5.93% in Garissa to 46.02% in Nyeri, with urban clusters achieving significantly higher coverage. Key predictors include bank access (financial inclusion), household crowding, and environmental factors (NO_2_ levels, precipitation). Wealth-based inequities (ECI = 0.044) were more pronounced than immunization-linked disparities (ECI = 0.025), highlighting financial barriers as the primary exclusionary factor.

**Conclusions:** This study underscores the need for targeted interventions, including mobile vaccination units, financial inclusion programs (e.g., M-Pesa subsidies), and integration of COVID-19 vaccines into routine immunization programs. Findings offer a replicable geospatial framework for low- and middle-income countries (LMICs), providing data-driven policy recommendations to enhance vaccine equity and pandemic preparedness. Addressing these disparities requires multisectoral approaches that integrate health system strengthening, financial accessibility, and climate resilience to ensure equitable vaccine distribution in vulnerable populations.

## 1 INTRODUCTION

The COVID-19 pandemic triggered an unprecedented global vaccination campaign, with over 13 billion doses administered by 2023, averting millions excess deaths and saving trillions in economic losses [1]. However, stark inequities emerged: by mid-2022, high-income countries had achieved 75-80% primary series coverage, while low-income nations struggled to reach even 10% [2]. Kenya exemplifies this disparity. Despite government campaigns and COVAX-supported initiatives, only 36% of Kenyans had received at least one vaccine dose by late 2023 [3], far below the African Union’s 60% target [4]. This shortfall reflects systemic failures that transcend individual choice: the majority of Kenya’s unvaccinated population resides in rural and arid regions, where healthcare deserts, seasonal floods, and longstanding distrust in health institutions create persistent barriers to access [5]. While mobile vaccination units and community outreach programs have made progress, these efforts remain fragmented and fail to account for the spatial clustering of vulnerabilities that perpetuate inequities.

Existing research on COVID-19 vaccine disparities has predominantly focused on individual-level factors such as age, education, and socioeconomic status, vaccination hesitancy, or single contextual barriers like travel time to clinics [5–7]. These approaches fail to capture the complex interplay of structural, geographic, and environmental determinants that define vaccine access in Kenya. For instance, wealth-based inequity measures often underestimate the true disparities in vaccine coverage, particularly when geographic and environmental factors, such as rurality and climate stressors, are considered [8]. Similarly, while Kenya’s routine childhood immunization rate (85%) far exceeds its COVID- 19 vaccination coverage [3], no study has explored whether using this existing health infrastructure could improve pandemic response efforts. The absence of spatially explicit, multidimensional analyses has left policymakers with limited actionable insights to break cycles of disadvantage and target resources effectively.

This study addresses these gaps by applying a geospatial epidemiological approach to examine the spatial determinants of COVID-19 vaccination disparities in Kenya. We investigate how socioeconomic deprivation, healthcare system fragmentation, and environmental stressors interact across geographic regions to shape vaccine uptake. Additionally, we assess whether historical immunization networks correlate with COVID-19 vaccine equity, providing insights into how Kenya’s existing health infrastructure could be optimized for future pandemic response strategies. Finally, we identify geographic clusters of compounded vulnerabilities that require urgent, prioritized intervention. Given that 75% of Kenya’s population resides in rural areas with fragmented health services, a spatial epidemiology approach is crucial to uncovering systemic drivers of inequity and designing targeted, data-driven solutions.

This study provides a spatially explicit, systemic understanding of vaccine disparities, equipping policymakers with the tools to design more equitable, evidence-based interventions in Kenya and beyond. By establishing a replicable model for implementing spatial epidemiology in low- and middle-income countries (LMICs), this research contributes to global efforts to reduce vaccine inequities and advance Sustainable Development Goal 3.8 (Universal Health Coverage) [9].

## 2 METHODS

This study employed a comprehensive multivariate spatial analysis framework to investigate the determinants of COVID-19 vaccination uptake across Kenya. By integrating socio-economic, environmental, and health data with advanced spatial and statistical techniques, we aimed to uncover spatial patterns and predictors of vaccination coverage. The analysis was structured into five main stages: (1) data aggregation and preprocessing, (2) correlation and spatial autocorrelation analysis, (3) spatial clustering using K-Means, (4) evaluation of covariate importance using Random Forest regression, and (5) Development Index construction and spatial associations.

### 2.1 Data Sources and Aggregation

#### 2.1.1 Data Sources

The study used multiple data sources. Socio-economic and health data were obtained from the Kenya Demographic and Health Survey (KDHS-2022) [3], which provided information from 42,300 households distributed across 1,692 georeferenced clusters (DHS-clusters). Variables included household wealth, sanitation facilities, roofing materials, handwashing facilities, cooking fuel type, bank account ownership, household size, age distribution, HIV prevalence, immunization rates, and COVID-19 testing and vaccination rates. Environmental data comprised climatic variables such as mean, maximum, and minimum temperature, and precipitation, obtained from BIOCLIM version 2.1. Data on extreme weather events (droughts and floods) and air pollution indicators like nitrogen dioxide (NO_2_) concentrations were extracted from spatial layers available in the Google Earth Engine (GEE) catalog. All environmental raster data were aligned with the socio- economic data by extracting raster values to the georeferenced cluster points.

The primary outcome variable in this study is the COVID-19 vaccination rate, which measures the proportion of eligible individuals within a household or cluster who have received at least one dose of the COVID-19 vaccine. This variable was derived from data collected in the KDHS-2022) and was computed as the ratio of vaccinated individuals to the total eligible population within each cluster. Eligibility was defined based on age and health criteria recommended for COVID-19 vaccination at the time of data collection. This metric serves as a critical measure for assessing disparities in vaccine uptake across socio-economic, environmental, and health dimensions.

#### 2.1.2 Data Preprocessing and Variable Construction

Categorical variables from the KDHS-2022 were recoded into ordinal variables to facilitate analysis. Similar categories were grouped, and new ordinal values were assigned using the *replace()* method from the Pandas library [10]. For instance, household wealth indices were coded from poorest (1) to richest (5). Data aggregation was performed to summarize household-level data at the cluster level using custom functions. For ordinal variables, medians were calculated to represent central tendencies. Means were computed for continuous variables like age and altitude. For binary variables, ratios or proportions were derived, such as the proportion of households with bank accounts. Modes were identified for categorical variables to represent the most common category. Tabs. S1 to S3 in Supplementary Materials summarize the variables included in the study.

Fig. 1 illustrates the conceptual model guiding this study. The model hypothesizes that COVID-19 vaccination uptake in Kenya is influenced by socio-economic factors (living conditions and financial access), health-related factors (healthcare access), environmental factors (air pollution and climatic variables), and demographic factors (child ratio and age distribution). Socio-economic factors such as the number of sleeping rooms and bank account ownership are expected to directly affect vaccination rates by influencing individuals’ ability to access vaccination services and adhere to public health recommendations. Health-related factors like immunization rates serve as indicators of trust in healthcare systems and may predict willingness to receive the COVID-19 vaccine. Environmental factors, including NO_2_ levels and precipitation, may impact both the spread of the virus and individuals’ health status, thereby affecting vaccination uptake. Demographic factors, such as the child ratio, can influence household priorities and resource allocation, potentially impacting vaccination decisions.

**Figure 1.**
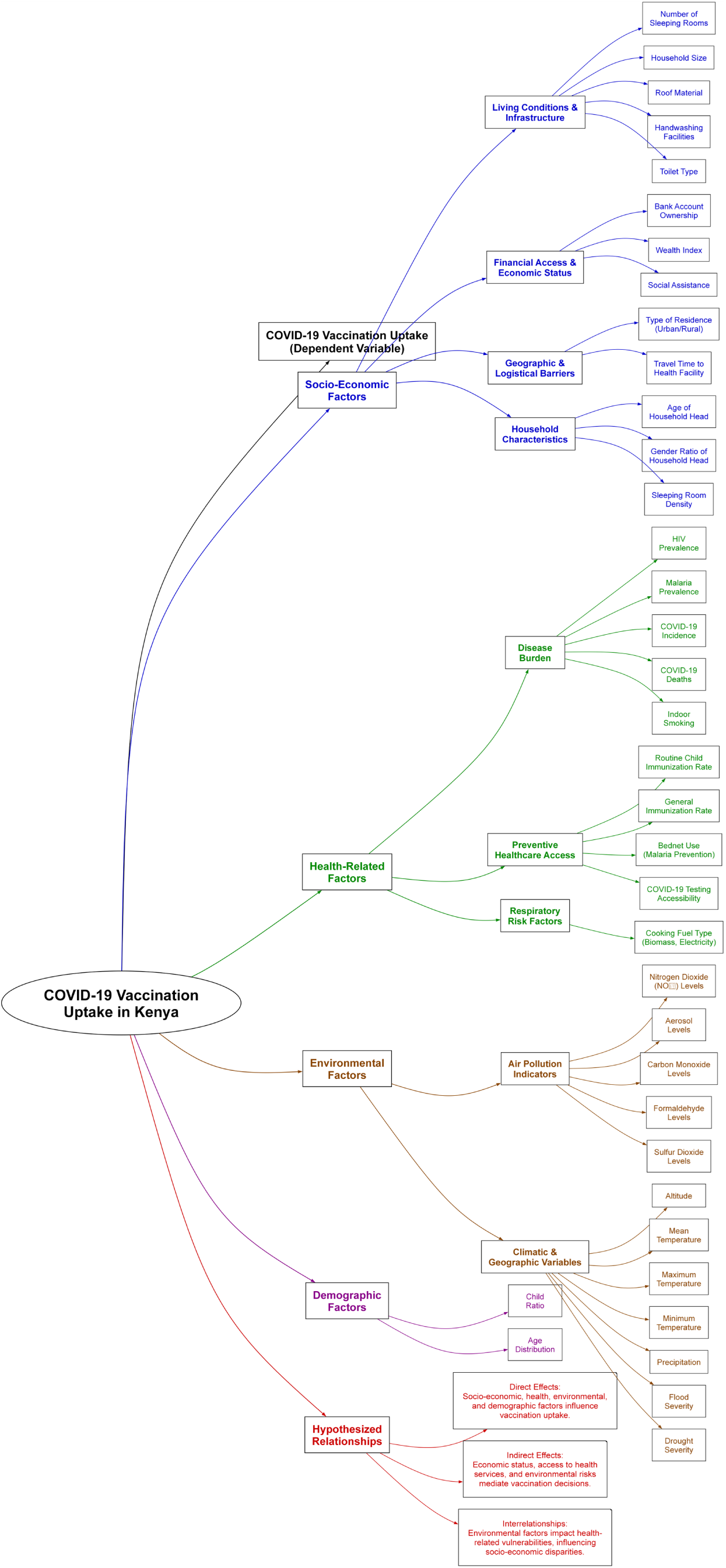
Conceptual model illustrating the hypothesized factors influencing COVID-19 vaccination uptake in Kenya.

### 2.2 Correlation and Spatial Autocorrelation Analysis

#### 2.2.1 Spearman Correlation Analysis

Spearman’s rank correlation coefficient was used to assess monotonic relationships between the dependent variables, COVID-19 testing rates and vaccination rates, and the explanatory variables. This non-parametric method is suitable for both continuous and ordinal data and captures non-linear relationships. Variables with correlation coefficients (𝜌) greater than 0.3 or less than -0.3 and a p-value less than 0.05 were considered significantly associated with the dependent variables.

#### 2.2.2 Spatial Weights Matrix Construction

A spatial weights matrix was constructed to model spatial relationships between clusters. The k-nearest neighbors method with 𝑘=12 was used to capture general regional patterns. The number of neighbors was chosen considering the total number of spatial entities and the regional scale of the dataset. The matrix was row-standardized to ensure each cluster’s weights summed to one, making influence proportional across clusters regardless of the number of neighbors. The weights module from the *pysal.lib* library was used for construction.

#### 2.2.3 Univariate Spatial Autocorrelation

Moran’s I statistic was calculated for the dependent variables and covariates identified as significant during the Spearman correlation analysis. Variables were considered to exhibit spatial dependence if their Moran’s I values were greater than 0.2 or less than -0.2, with a p-value less than 0.05. Moran’s I statistic was implemented using the *esda* module from *pysal.lib*.

#### 2.2.4 Bivariate Spatial Autocorrelation

Bivariate Moran’s I was calculated to assess the spatial relationship between vaccination rates and each significant covariate during the univariate Moran’s I analysis. Covariates were considered to exhibit spatial dependence if their Bivariate Moran’s I values were greater than 0.1 or less than -0.1, with a p-value less than 0.05. This analysis was implemented through the *esda.Moran BV* function from *pysal.lib*.

#### 2.2.5 Spatial Lag Model (SLM)

A Spatial Lag Model was fitted to account for mutivariate spatial dependencies in vaccination rates and significant covariates during the bivariate Moran’s I analysis. The SLM considered both the influence of covariates and the spatial effect of vaccination rates in neighboring clusters. The model was implemented using the *ML Lag* function from the *spreg* module in the *pysal* library.

### 2.3 Spatial K-Means Clustering

#### 2.3.1 Data Preparation

Significant covariates from the SLM were standardized using the *StandardScaler* from the *sklearn.preprocessing* module to ensure equal contribution to clustering. Standardization centered the variables around zero and scaled them by their standard deviation. Geographic coordinates (latitude and longitude) were appended to the dataset to incorporate spatial proximity into the clustering process.

#### 2.3.2 Determination of Optimal Clusters

The optimal number of K-Means clusters was determined using the Elbow Method. K-Means clustering was performed for 𝑘 values ranging from 1 to 15 using *n clusters=k* to the standardized covariates from *kmeans.fit()* function. Inertia (sum of squared distances to cluster centers) was calculated with *kmeans.inertia* and plotted against 𝑘.

#### 2.3.3 K-Means Clustering Implementation

The K-Means clustering method was applied to the dataset to identify socio-geographic groupings of clusters based on shared characteristics. The clustering model was trained on a subset of standardized covariates that were significantly associated with vaccination rates in previous analyses. These variables included handwashing facilities, bank access, sleeping room density, annual precipitation, NO_2_ levels, household size, and child ratio. Geographic coordinates (latitude and longitude) were also appended to ensure spatial awareness in the clustering process.

K-Means clustering was performed with 𝑘 = 7 using the *KMeans* function from the *scikit-learn* library. Each cluster was assigned to one of seven clusters based on similarity in standardized covariates and spatial proximity. The clustering classification was incorporated into the dataset, and results were exported as spatial layers with point geometry based on the latitude and longitude data for mapping and further analysis.

### 2.4 Random Forest Analysis

#### 2.4.1 Model Preparation

The Random Forest regression model was used to predict vaccination rates and assess covariate importance. The dataset was split into training (70%) and testing (30%) subsets using the *train test split* function from the *sklearn.model selection* module, ensuring independence in model evaluation and reducing the risk of overfitting. The feature set included the significant covariates identified in the Spearman correlation analysis and the cluster labels from the K-Means clustering.

#### 2.4.2 Hyperparameter Optimization

Hyperparameters were optimized using grid search with five-fold cross-validation implemented through the *GridSearchCV* function. The parameters tuned included the number of trees (*n estimators*), maximum depth of the trees (*max depth*), minimum number of samples required to split an internal node (*min samples split*), minimum number of samples required to be at a leaf node (*min samples leaf* ), and whether bootstrap samples were used when building trees (*bootstrap*). The best model was retrieved using *grid search.best estimator* .

#### 2.4.3 Model Evaluation

Model performance was assessed on the test set using the optimized model. Metrics included Mean Squared Error (MSE) and the coefficient of determination (𝑅^2^). The *predict()* function was used to perform this calculation.

### 2.5 Development Index Construction

To synthesize the multidimensional factors influencing vaccination rates, a Development Index (DI) was constructed as a weighted sum of key socioeconomic, healthcare, environmental, and demographic components to synthesize the dimensions affecting vaccination rates in each of the DHS-clusters. The index was derived from the Random Forest variable importance analysis, with weights assigned based on each factor’s relative contribution to vaccination rate prediction. The final index was normalized to a 0–100 scale, with higher values indicating more favorable conditions for vaccination uptake. Covariate importance was calculated using the *feature importances* attribute of the Random Forest model, which returns the normalized relative contribution of each variable. The index was scaled from 0 to 100 using the formula:

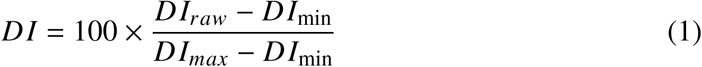

This composite index provided a unified metric to assess regional disparities and their association with vaccination rates.

### 2.6 Spatial and household Equity Analysis with Concentration Index

Bivariate maps were created to visualize the spatial relationship between COVID-19 vaccination rates and key covariates, including child immunization coverage and wealth index. DHS-cluster-level data for COVID-19 vaccination rates were combined with the corresponding covariate values, and each DHS-cluster was assigned a color gradient representing the combination of high and low values for both variables. Darker shades indicated clusters with high values for both variables, while lighter shades represented low values. Intermediate shades reflected mixed patterns, with one variable high and the other low.

To assess socioeconomic and health system inequities in COVID-19 vaccination uptake at household level, we calculated the Erreygers Concentration Index (ECI) [11] for two dimensions:

1. Wealth-based inequality: Households ranked by wealth index (1 = poorest to 5 = richest).
2. Immunization-linked inequality: Clusters ranked by childhood immunization rates (continuous scale).

The ECI was computed as:

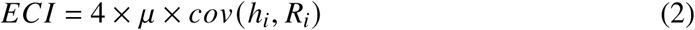

where 𝜇 is the mean vaccination rate, ℎ_𝑖_ is the vaccination status of household 𝑖, and 𝑅_𝑖_ is fractional rank of the equity variable (wealth or immunization). Clusters were sorted by equity variables, and concentration curves were plotted to visualize cumulative vaccination proportions against cumulative population shares. Positive ECI values indicate pro-rich/pro-immunization inequality; negative values signal pro-poor/compensatory patterns.

### 2.7 Technical Implementation

All analyses were implemented using Python version 3.11.10 [12]. The primary libraries included Pandas (v2.2.3) for data manipulation, NumPy (v2.0.2) for numerical computations, scikit-learn (v1.5.2) for machine learning algorithms (e.g., *StandardScaler, KMeans, RandomForestRegressor, GridSearchCV*), PySAL (v4.12.1) for spatial analysis (e.g., *spatial weights, Moran’s I*), and Matplotlib (v3.9.2) and Seaborn (v0.13.2) for data visualization. Household-level data were aggregated at the cluster level using appropriate protocols: arithmetic means for continuous variables (*df.groupby(’cluster’).mean()* in Pandas), medians for ordinal variables (*df.groupby(’cluster’).median()*), and proportions for binary variables (*df.groupby(’cluster’).apply(lambda x: x.mean())*). Additionally, categorical variables were aggregated using modal frequency (*df.groupby(’cluster’).agg(lambda x: x.mode()*[0])). These transformations ensured consistency across different data types and allowed for meaningful spatial comparisons. Statistical analysis was conducted in R using the *dplyr* and *ggplot2* packages [13]. Maps were generated using ArcGIS Pro [14].

## 3 RESULTS

### 3.1 Descriptive Statistics and Spatial Distribution of Vaccination Coverage

The national average COVID-19 vaccination rate across Kenya was 27%. Urban areas reported a higher vaccination coverage (30.22%) compared to rural areas (25.36%) (p ¡ 0.05). Significant disparities were observed between counties, with Nyeri displaying the highest vaccination rate (46.02%) and Garissa the lowest (5.93%).

### 3.2 Exploratory Analysis and Spatial Correlation

Tab. 1 summarizes the results of exploratory analyses, spatial correlation assessments, and covariate importance evaluations regarding COVID-19 vaccination rates. The Spearman correlation analysis revealed significant associations between vaccination rates and multiple socioeconomic, health, and environmental factors. Wealth index, bank access, and handwashing facilities were positively correlated with vaccination rates, whereas larger household size, a higher proportion of children, and lower accessibility to health services exhibited negative correlations. Environmental variables such as temperature and precipitation also displayed associations with vaccine uptake, suggesting the influence of climate-related barriers on access to healthcare.

**Table 1.** Spatial Correlations and Covariate Importance for outcome variables.

| Variable | Testing ($\rho$) | Vaccination ($\rho$) | Univariate Moran's I ($p$) | Bivariate Moran's I ($p$) | SLM Coef. ($p$) | Cov. Imp. |
| --- | --- | --- | --- | --- | --- | --- |
| **Socioeconomic** |  |  |  |  |  |  |
| Wealth Index | **0.520\*\*** | **0.490\*\*** | **0.424\*\* ($< 0.05$)** | **0.318\*\* ($< 0.05$)** | -0.003 ($> 0.05$) | 0.013 |
| Bank Access | **0.432\*\*** | **0.553\*\*** | **0.436\*\* ($< 0.05$)** | **0.357\*\* ($< 0.05$)** | **0.118\*\* ($< 0.05$)** | 0.115 |
| Household Size | **-0.427\*\*** | **-0.495\*\*** | **0.369\*\* ($< 0.05$)** | **-0.327\*\* ($< 0.05$)** | **-0.001\*\* ($< 0.05$)** | 0.068 |
| Age of Household Head | **-0.319\*\*** | -0.024 | **0.265\*\* ($< 0.05$)** | 0.019 ($> 0.05$) | | 0.028 |
| Child Ratio | -0.176 | **-0.438\*\*** | **0.300\*\* ($< 0.05$)** | **-0.326\*\* ($< 0.05$)** | **-0.259\*\* ($< 0.05$)** | 0.031 |
| Roof Material | 0.257 | **0.358\*\*** | **0.347\*\* ($< 0.05$)** | **0.281\*\* ($< 0.05$)** | -0.002 ($> 0.05$) | 0.001 |
| Handwashing Place | 0.247 | **0.377\*\*** | **0.480\*\* ($< 0.05$)** | **0.277\*\* ($< 0.05$)** | **0.013\*\* ($< 0.05$)** | 0.007 |
| Water Source | 0.290 | 0.190 |  |  |  |  |
| Social Assistance | -0.220 | -0.107 |  |  |  |  |
| Number of sleeping rooms per household | 0.255 | **0.604\*\*** | **0.565\*\* ($< 0.05$)** | **0.437\*\* ($< 0.05$)** | **0.171\*\* ($< 0.05$)** | 0.328 |
| Gender Ratio | 0.062 | -0.017 |  |  |  |  |
| Toilet Type | **0.399\*\*** | **0.368\*\*** | **0.327\*\* ($< 0.05$)** | | 0.002 ($< 0.05$) | |
| Travel Time by Vehicle | **-0.328\*\*** | **-0.330\*\*** | 0.075 ($< 0.05$) | | 0.018 ($< 0.05$) | |
| Travel Time Without Vehicle | -0.287 | **-0.318\*\*** | **0.223\*\* ($< 0.05$)** | **-0.262\*\* ($< 0.05$)** | 0.000 ($> 0.05$) | 0.013 |
| **Health** |  |  |  |  |  |  |
| HIV Prevalence | 0.270 | **0.453\*\*** | 0.142 ($< 0.05$) | **0.104\*\* ($< 0.05$)** | 0.000 ($> 0.05$) | 0.026 |
| Malaria Prevalence | -0.038 | 0.078 |  |  |  |  |
| Child Immunization Rate | 0.058 | **0.378\*\*** | **0.853\*\* ($< 0.05$)** | **0.417\*\* ($< 0.05$)** | 0.015 ($> 0.05$) | 0.032 |
| Bednet Use | -0.059 | 0.006 |  |  |  |  |
| Indoor Smoking | -0.024 | 0.006 |  |  |  |  |
| COVID-19 Incidence | **0.302\*\*** | 0.287 | 0.076 ($< 0.05$) | | 0.013 ($< 0.05$) | |
| COVID-19 Deaths | 0.042 | 0.017 |  |  |  |  |
| Cooking Fuel | **0.510\*\*** | 0.299 | **0.404\*\* ($< 0.05$)** | **0.191\*\* ($< 0.05$)** | -0.001 ($> 0.05$) | 0.005 |
| **Environmental** |  |  |  |  |  |  |
| Altitude | 0.123 | **0.350\*\*** | **0.932\*\* ($< 0.05$)** | **0.399\*\* ($< 0.05$)** | 0.000 ($> 0.05$) | 0.069 |
| Mean Temperature | -0.142 | **-0.392\*\*** | -0.002 ($< 0.05$) | | 0.039 ($< 0.05$) | |
| Maximum Temperature | -0.119 | **-0.379\*\*** | -0.002 ($< 0.05$) | | 0.017 ($< 0.05$) | |
| Minimum Temperature | -0.150 | **-0.386\*\*** | -0.003 ($< 0.05$) | | | 0.073 |
| Precipitation | 0.086 | **0.388\*\*** | **0.680\*\* ($< 0.05$)** | **0.348\*\* ($< 0.05$)** | **0.000\*\* ($< 0.05$)** | 0.021 |
| Flood Severity | -0.019 | -0.123 |  |  |  |  |
| Nitrogen Dioxide (NO<sub>2</sub>) Levels | **0.316\*\*** | **0.406\*\*** | **0.974\*\* ($< 0.05$)** | **0.316\*\* ($< 0.05$)** | **2450.278\*\* ($< 0.05$)** | 0.073 |
| Drought Severity | 0.153 | 0.135 |  |  |  |  |
| Aerosols Levels | -0.039 | -0.299 |  |  |  |  |
| Carbon Monoxide Levels | -0.092 | -0.216 |  |  |  |  |
| Formaldehyde Levels | 0.128 | 0.172 |  |  |  |  |
| Sulfur Dioxide Levels | 0.147 | 0.179 |  |  |  |  |

Spatial autocorrelation analysis using Moran’s I identified strong clustering in vaccination rates, with an observed Moran’s I of 0.531 (p < 0.05), confirming that vaccination coverage is not randomly distributed but exhibits spatial dependence. Conversely, COVID-19 testing rates showed weak spatial dependence (Moran’s I = 0.060, p < 0.05), leading to their exclusion from further spatial modeling. Among the independent variables, wealth, bank access, sleeping room density, and nitrogen dioxide levels exhibited strong positive spatial autocorrelation, while household size and child ratio demonstrated negative spatial autocorrelation, indicating clustered patterns of disadvantage.

Bivariate Moran’s I analysis assessed spatial associations between vaccination rates and key covariates. Significant positive spatial relationships were found for wealth index (I = 0.318, p < 0.05), handwashing facilities (I = 0.277, p < 0.05), and bank access (I = 0.356, p ¡<0.05), suggesting that vaccination rates tend to be higher in areas with better infrastructure and financial access. Conversely, variables such as household size (I = -0.327, p < 0.05) and child ratio (I = -0.326, p < 0.05) exhibited negative spatial correlations, indicating that these demographic factors may serve as barriers to vaccination uptake.

### 3.3 Spatial Dependence and Structural Determinants of Vaccination

To further disentangle the spatial structure of vaccine coverage, a Spatial Lag Model (SLM) was fitted to account for the influence of neighboring regions on local vaccination rates. The model achieved a pseudo 𝑅^2^ of 0.6333 and a spatial pseudo 𝑅^2^ of 0.5651, indicating that spatial factors play a crucial role in shaping vaccination disparities. The results showed that financial access (p < 0.001) and sleeping room density (p < 0.001) had significant positive effects on vaccination rates, while child dependency ratio (p < 0.001) and household size (p < 0.001) negatively impacted vaccine uptake. Environmental variables, particularly nitrogen dioxide (NO_2_) levels (p < 0.001), also exhibited strong spatial effects, suggesting that pollution levels may contribute to disparities in vaccination rates.

### 3.4 Spatial Clustering of Vaccination Coverage

Spatial K-Means clustering was applied to identify geographic patterns of vaccine coverage disparities. Standardized socioeconomic, health, and environmental variables were incorporated alongside geographic coordinates to enhance the clustering analysis. The optimal number of clusters was determined using the elbow method, which identified seven distinct clusters (Fig. S1 in Supplementary Materials). Cluster membership revealed pronounced urban-rural disparities, with urban areas demonstrating significantly higher vaccination rates than rural regions. Specifically, clusters with the highest vaccination rates exhibited better living conditions, greater financial access, and lower dependency ratios, while clusters with the lowest vaccination rates were characterized by larger household sizes, lower bank access, and poor environmental conditions.

### 3.5 Predictive Modeling: Random Forest Analysis

A Random Forest regression model was developed to quantify the relative importance of covariates in predicting vaccination rates. The optimized model, selected through cross-validation, consisted of 200 decision trees with a maximum depth of 10, achieving a mean squared error (MSE) of 0.0076 and an 𝑅^2^ of 0.62. These results indicate that the model captures a substantial portion of the variability in vaccination rates, albeit with some remaining unexplained variance.

Feature importance analysis (Fig. 2) highlighted sleeping room density and bank access as the most influential predictors of vaccination rates. Other significant predictors included altitude, child ratio, and nitrogen dioxide (NO_2_) levels, which collectively accounted for a large proportion of model variance. Notably, socioeconomic factors such as wealth index and roof material exhibited relatively low predictive importance when compared to environmental and demographic factors, underscoring the complex, multidimensional nature of vaccine access disparities.

**Figure 2.**
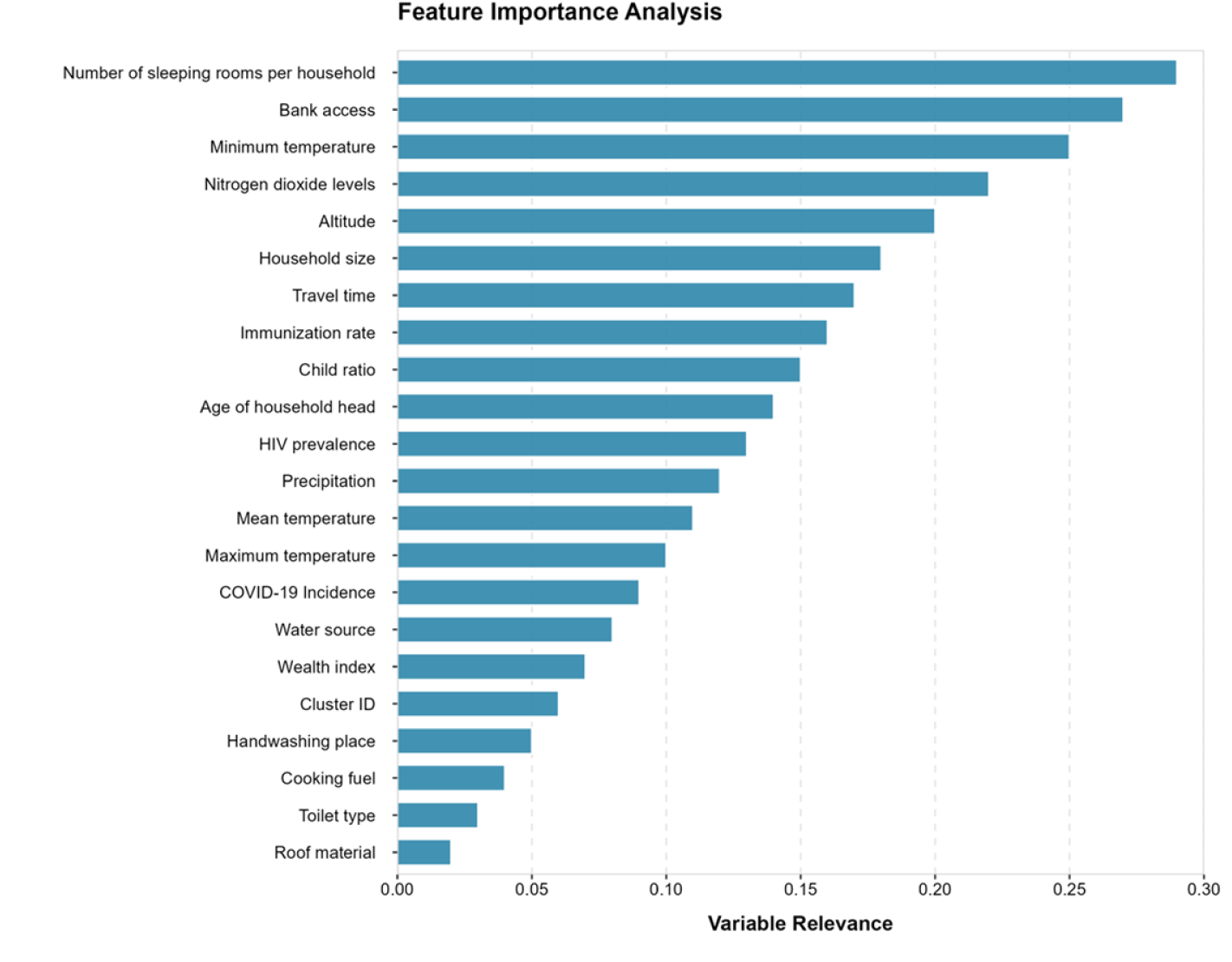
Relative importance of covariates from Random Forest model. This horizontal bar plot displays the relative relevance of covariates from a Random Forest model predicting COVID-19 vaccination outcomes. Variables are ordered vertically from highest to lowest relevance, with number of sleeping rooms per household (0.30) emerging as the most influential predictor and roof material (0.02) showing the minimal impact. The relevance metric (x-axis) ranges from 0.00 to 0.30, representing each variable’s normalized importance score calculated through mean decrease in impurity. Notable patterns include strong environmental predictors (tmin, no2, tmax), moderate socioeconomic factors (access to bank account, wealth index), and weaker infrastructure-related variables (toilet, cooking material). All relevance scores were derived from 500 trees with out-of-bag error estimation.

### 3.6 Development Index (DI)

The DI was calculated in each DHS-cluster by standardizing and weighting the nine key variables identified in the random forest analysis across five components: living conditions, financial access, healthcare indicators, environmental factors, and demographics. Living conditions, represented by the number of sleeping rooms per household member and household size, contributed the highest weight (39.53%) based on Random Forest feature importance. Financial access, including bank ownership and wealth index, accounted for 12.79%, while healthcare indicators such as child immunization coverage and HIV prevalence contributed 5.83%. Environmental factors, including nitrogen dioxide (NO_2_) levels and handwashing facilities, accounted for 8.00%, and demographic factors, represented by child ratio, contributed 3.09%. The raw composite DI was calculated as the weighted sum of these standardized components and rescaled to a 0–100 range using Eq. (1). Higher DI scores were observed in urban clusters, reflecting better financial inclusion, improved living conditions, and higher immunization rates. Conversely, rural clusters, particularly Rural Remote (Cluster 5) and Rural Trans (Cluster 2), exhibited lower DI scores, indicating limited access to infrastructure, healthcare, and financial resources.

The DI revealed substantial geographic disparities, with the lowest-scoring regions aligning with clusters identified as having low vaccination rates in the spatial K-Means clustering analysis (Fig. 3). The spatial clustering analysis identified seven clusters with distinct combinations of socioeconomic, infrastructural, and geographic characteristics influencing COVID-19 vaccination coverage. Vaccination rates DI scores exhibited a clear gradient, increasing from rural to urban clusters. Rural-Remote clusters (Cluster 5), located predominantly in northern and northeastern Kenya, had one of the lowest vaccination rates, averaging 7.84%, and the lowest DI scores, reflecting limited healthcare access, financial inclusion, and adverse environmental conditions. Rural Trans (Cluster 2), situated in semiarid transition zones, demonstrated very low vaccination rates and modest improvements in financial inclusion. Agriculturally productive regions, categorized as Rural Dev (Cluster 1), had moderate vaccination rates, supported by better child immunization coverage and living conditions.

**Figure 3.**
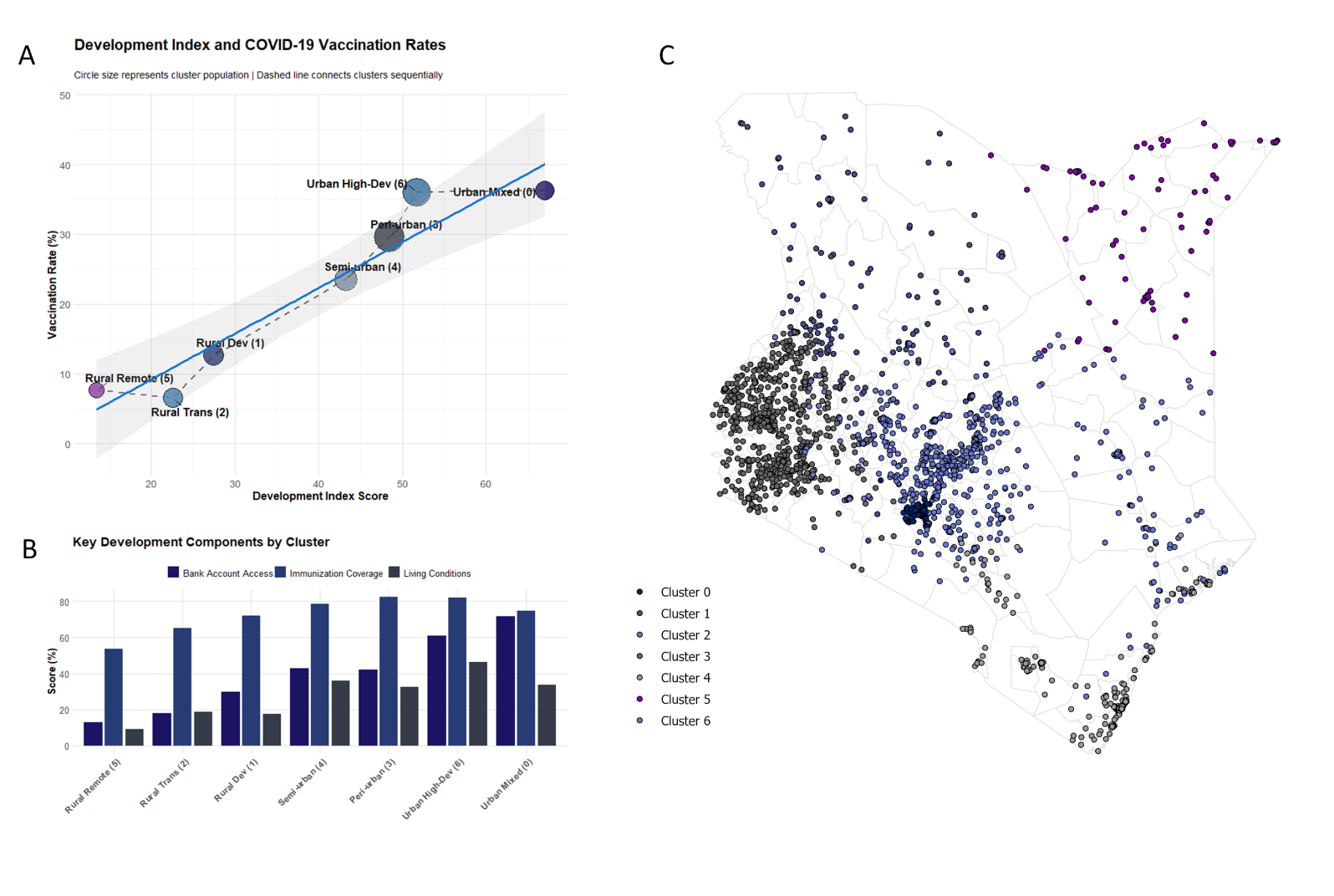
Development Index, COVID-19 Vaccination Rates, and Cluster-Level Characteristics in Kenya. (A) Scatterplot showing the relationship between the Development Index (DI) and COVID-19 vaccination rates across seven clusters identified through K-means clustering. Circle size represents cluster population, and the dashed line connects clusters sequentially based on DI scores. A positive linear trend is observed, with higher vaccination rates corresponding to higher DI scores. Clusters are labeled by their development category: Rural Remote (Cluster 5), Rural Trans (Cluster 2), Rural Dev (Cluster 1), Semi-urban (Cluster 4), Peri-urban (Cluster 3), Urban High-Dev (Cluster 6), and Urban Mixed (Cluster 0). (B) Bar plot presenting the scores for key DI components, Bank Account Access, Immunization Coverage, and Living Conditions, by cluster. Urban High-Dev and Urban Mixed clusters exhibit the highest scores across all components, while Rural Remote clusters show the lowest values, highlighting disparities in development and healthcare access. (C) Geographic distribution of K-mean clusters across Kenya, illustrating the spatial clustering of development categories. Urban clusters are concentrated in major cities and peri-urban regions, while rural clusters dominate northern and northeastern areas, characterized by lower DI scores and vaccination rates.

Semi-urban (Cluster 4) and Peri-urban (Cluster 3) clusters, near smaller towns and urban fringes, exhibited intermediate vaccination rates, benefiting from partial access to urban infrastructure. Urban High-Dev clusters (Cluster 6), located in major cities such as Nairobi and Mombasa, achieved the highest vaccination rates and DI scores due to strong financial inclusion, healthcare access, and living conditions. Urban Mixed clusters (Cluster 0) also achieved high vaccination rates but displayed variability in DI components.

The relationship between the DI and vaccination rates, shown in Fig. 3A, revealed a positive linear trend. Urban clusters achieved higher vaccination rates, exceeding 31.51%, while rural clusters such as Rural Trans and Rural Remote averaged 6.56% and 7.84% respectively. Fig. 3B decomposes the DI into Bank Account Access, Immunization Coverage, and Living Conditions (measured by sleeping rooms per household member). Urban clusters consistently outperformed rural clusters across all components, with Urban High-Dev clusters achieving particularly high financial inclusion. Child immunization coverage was higher in rural clusters, but logistical challenges and weak healthcare systems limited their COVID-19 vaccination rates.

### 3.7 Spatial and household Equity Analysis

The analysis of COVID-19 vaccination rates, routine child immunization coverage, and wealth levels revealed significant spatial and household-level disparities. The bivariate map (Fig. 4A) showed that regions with high COVID-19 vaccination rates aligned spatially with areas of high child immunization coverage, particularly in urban centers and agriculturally productive regions. In contrast, regions with low vaccination and immunization rates were concentrated in northern and northeastern Kenya. The scatterplot (Fig. 4B) highlighted a positive association between COVID-19 vaccination and child immunization rates, with clusters exhibiting both high vaccination and immunization rates located in the upper-right quadrant. Clusters deviating from this trend suggested additional barriers, such as logistical challenges.

**Figure 4.**
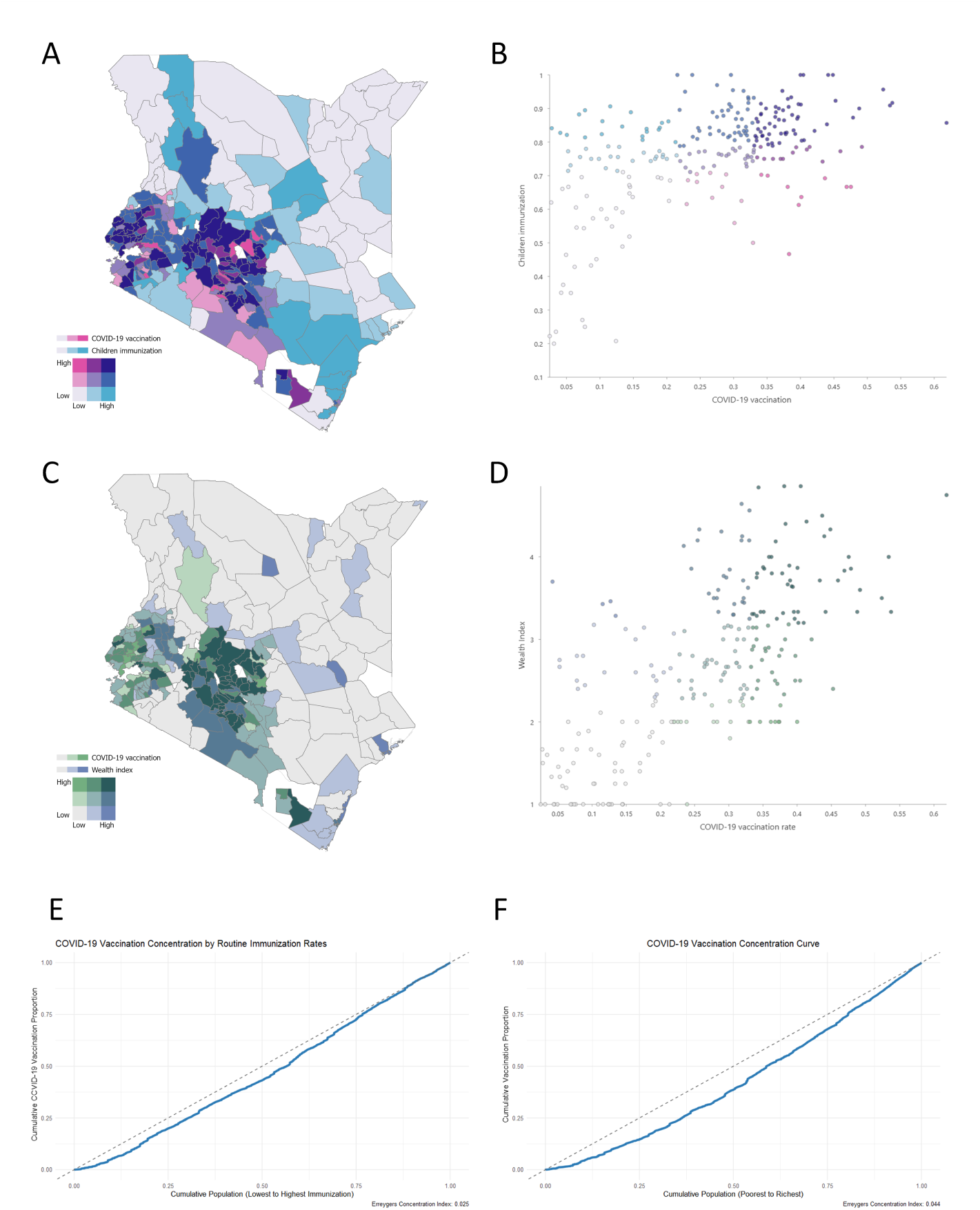
Spatial, Household-Level, and Equity Analyses of COVID-19 Vaccination Coverage in Kenya. (A) Bivariate map showing the spatial distribution of COVID-19 vaccination rates and child immunization coverage. Dark purple regions indicate areas with high vaccination and immunization rates, while light blue regions represent areas with low rates for both variables. Spatial overlap suggests that regions with strong routine immunization programs also exhibit higher COVID-19 vaccination rates, particularly in urban and agriculturally productive regions. (B) Scatterplot illustrating the positive association between COVID-19 vaccination rates and child immunization coverage at the cluster level. Clusters in the upper-right quadrant exhibit both high vaccination and immunization rates, reflecting areas with robust healthcare systems, while clusters in the lower-left quadrant represent regions with overlapping challenges. (C) Bivariate map showing the spatial relationship between COVID-19 vaccination rates and wealth index. Dark green regions indicate areas with high vaccination rates and wealth, while light green areas represent low rates for both variables. High-wealth, high-vaccination regions are concentrated in urban centers, while low-wealth, low-vaccination regions dominate rural and remote areas. (D) Scatterplot showing the positive association between COVID-19 vaccination rates and wealth index. While wealthier clusters tend to achieve higher vaccination rates, the dispersed pattern highlights variability due to other factors such as healthcare access and geographic barriers. (E) Concentration curve for COVID-19 vaccination rates based on child immunization coverage, yielding an Erreygers Concentration Index (ECI) of 0.025. The curve demonstrates a moderate pro-rich distribution, with higher vaccination rates favoring regions with better immunization infrastructure. (F) Concentration curve for COVID-19 vaccination rates based on wealth index, with an ECI of 0.044. The curve reflects more pronounced inequities, with wealthier regions disproportionately benefiting from vaccine distribution.

The spatial distribution of vaccination rates and wealth levels (Fig. 4C) showed strong alignment in high-wealth regions, particularly urban centers, while low-wealth regions, mostly rural and remote, had lower vaccination rates. The scatterplot (Fig. 4D) revealed a positive but dispersed association between vaccination rates and wealth, with wealthier clusters achieving higher vaccination rates and variability driven by non-financial barriers such as geographic accessibility.

The inequities in vaccine coverage were quantified using the Erreygers Concentration Index (Fig. 4E and Fig. 4F). The ECI for routine immunization coverage was 0.025, indicating a moderate pro-rich distribution of COVID-19 vaccinations, reflecting reliance on pre-existing immunization systems. The ECI for wealth was higher, at 0.044, indicating more pronounced inequities driven by economic disparities. The higher ECI for wealth highlights the significant role of financial disparities in shaping vaccine access beyond the influence of immunization infrastructure.

## 4 DISCUSSION

This study reveals profound and systematic inequities in COVID-19 vaccination coverage across Kenya, underscoring that a modest national average of 27% masks vastly divergent realities. Urban areas, including Nyeri, achieve substantially higher vaccination rates of up to 46%, while rural and arid regions such as Garissa barely reach about 6%. By merging geospatial clustering methods with machine learning and a composite DI, we show that these disparities are not incidental outcomes of personal decisions. Instead, they reflect entrenched structural disadvantages in healthcare infrastructure, financial inclusion, and environmental conditions. Notably, K-Means clustering identified seven distinct spatial groupings, revealing how an interconnected set of variables, ranging from the density of sleeping rooms in households to bank account ownership and exposure to seasonal floods, shapes the likelihood of receiving a COVID-19 vaccine. In many remote or arid parts of the country, coverage remains below 8%, pointing to significant logistical barriers and systemic neglect. At the same time, regions with higher nitrogen dioxide (NO_2_) levels, typically a proxy for urban development, displayed better vaccination rates due to potentially stronger healthcare networks and better resource availability.

These findings resonate with analogous inequities in other LMICs, indicating that financial hardship, fragmented healthcare systems, and challenging environments converge to create what can be described as ’vaccine deserts.’ By making these inequalities visible, our study offers a fresh perspective on the underlying drivers of low vaccination coverage, reinforcing global health discourses that emphasize how place-based disadvantage, rather than mere hesitancy, drives unequal access to life-saving health interventions [15]. The broader implications extend to any resource-constrained setting where the interplay of poverty, environmental shocks, and uneven service provision hinders efforts to control infectious diseases.

Interpretation of our results underscores how socioeconomic constraints, environmental hazards, and uneven health systems jointly dictate vaccine access in Kenya. The Random Forest analysis confirms that financial inclusion, reflected by access to formal banking, is a decisive factor, likely because banked households are better positioned to handle indirect vaccination costs such as transport and lost wages. Meanwhile, high sleeping room density emerges as a compelling proxy for overcrowded living conditions, associating strongly with low uptake in regions already facing pronounced poverty. These structural vulnerabilities are magnified by environmental disruptions: seasonal floods and complex terrain limit the mobility of healthcare workers, disrupt cold-chain maintenance, and render certain communities virtually unreachable during rainy seasons.

Contrary to assumptions that air pollution invariably signals poor health outcomes [16, 17], our data show a more complex relationship, where higher nitrogen dioxide (NO_2_) levels correlate with enhanced vaccination coverage, presumably reflecting urban infrastructure advantages that overshadow pollution’s detrimental effects. Similar patterns have been observed in countries like Nigeria [18] and Bangladesh [19], where major cities accumulate health services, specialized healthcare workers, and donor-funded projects, thus achieving superior vaccine outcomes compared with remote, environmentally challenging locales. Together, these insights call attention to the critical role of place-based development strategies that tackle the intersecting domains of infrastructure, climate resilience, and financial empowerment. In the Kenyan context, bridging the gulf between urban hubs and marginalized peripheries necessitates a coordinated response in which multiple sectors, ranging from transport and agriculture to digital finance, converge with public health policy. This integrated vision resonates with Africa’s Agenda 2063, which frames inclusive economic growth and robust health systems as mutually reinforcing pathways toward regional stability [20, 21]. Our study thus adds empirical weight to calls for policies that are no longer framed in single domains but rather align with a broader, equity-focused strategy for advancing public health security.

Translating the findings of our study into actionable policy requires addressing the social, infrastructural, and climatic barriers that perpetuate low vaccination rates in marginalized settings. Scaling up mobile vaccination programs is a vital step, especially for remote northern counties where traditional, facility-based approaches remain insufficient. Mobile teams, synchronized with post-flood recovery periods, can reduce both cost and travel time, as demonstrated in Uganda’s door-to-door campaigns that boosted rural vaccination coverage [22, 23]. However, mobile units alone cannot overcome the underlying issue of financial exclusion, which emerged as a principal predictor in our analyses. Collaborations with fintech platforms such as M-Pesa could allow governments to offer transport stipends or partial wage compensation to individuals missing work to get vaccinated, thereby removing a critical barrier for low-income households [24].

Another immediate opportunity lies in integrating COVID-19 vaccination with Kenya’s well-established childhood immunization programs that already reach around 85% coverage nationwide [3]. By capitalizing on existing cold-chain networks and community trust, health authorities could expedite vaccine delivery to the hardest-hit zones, mirroring Rwanda’s successful alignment of COVID-19 campaigns with child immunization outreach [25, 26]. Beyond these immediate measures, our study underscores the necessity of climate-resilient health infrastructure, particularly in flood-prone and arid counties. Investments in durable roads, drone-based vaccine deliveries, and decentralized storage facilities will not only improve emergency preparedness but also enhance routine health services in historically neglected communities. While these interventions may appear costly, evidence suggests that their long-term returns in saved medical expenditures and improved health outcomes are substantial. Indeed, each percentage increase in coverage may avert significant treatment costs downstream and mitigate the economic fallout of future pandemics. Taken together, these approaches endorse a multisectoral strategy that prioritizes equity, anticipating a broader transformation in how public health systems operate under conditions of climate and financial stress.

A key implementation of our research lies in its multifaceted methodology, integrating spatial autocorrelation measures, K-Means clustering, and Random Forest regression to capture intricate dynamics that traditional analyses might overlook. By linking socioeconomic indicators, environmental hazards, and health infrastructure metrics, we uncovered how overlapping deficits can entrench low vaccination coverage. The DI further synthesizes these domains into a single, interpretable metric, enabling policymakers to identify emergent ’hotspots’ of vulnerability. Real-time monitoring systems that combine routine health data with climate forecasting would offer a dynamic lens on evolving coverage gaps, by demonstrating that a composite DI and spatial clustering techniques can pinpoint pockets of vulnerability, our approach offers a universal toolkit. Health systems worldwide can use similar data-driven, geospatial methods to assess local vaccination challenges and allocate resources more equitably. Furthermore, developing real-time spatial monitoring systems is a key priority for improving vaccine delivery in crisis settings. GIS-based dashboards that link health facility data with vaccine stock levels and coverage metrics could enable public health authorities to monitor coverage gaps and respond proactively to emerging challenges, such as vaccine stockouts or climatic disruptions that impede access. Finally, expanding the DI framework to other LMICs would test its adaptability in regions grappling with similarly complex constraints, thereby creating a global repository of comparative insights. Taken together, these paths underscore the imperative of continuous methodological refinement and cross-sectoral data sharing, which could ultimately strengthen pandemic preparedness beyond the immediate challenges posed by COVID-19.

While our study provides valuable insights into spatial vaccination disparities, several important limitations warrant discussion. First, our reliance on cross-sectional data from the KDHS means we cannot capture temporal changes in vaccination coverage or assess causal relationships. DHS data, while nationally representative, has inherent limitations: surveys are typically conducted every 3-5 years, potentially missing rapid changes in health behaviors during crisis periods like the COVID-19 pandemic. Furthermore, the cluster-based sampling design of DHS may underrepresent highly mobile populations or those living in sparsely populated areas, particularly relevant in Kenya’s arid and semi-arid regions where nomadic communities reside. Another important limitation is our dependence on self-reported vaccination status, which may introduce recall bias and social desirability bias. Respondents might overreport vaccination to appear compliant with public health recommendations or underreport due to stigma or political beliefs.

Without verification against immunization records, we cannot assess the accuracy of these self-reported measures. Similarly, other key variables such as household wealth indicators, and health behaviors rely on self-reporting, potentially affecting the reliability of our socioeconomic analyses.

Likewise, the spatial resolution of our analysis was constrained by DHS confidentiality protocols, which randomly displace cluster coordinates to protect participant privacy. This displacement (up to 2 kilometers for urban clusters and 5 kilometers for rural clusters) may affect the precision of our spatial statistics and environmental exposure estimates. Additionally, we lacked data on several potentially important factors, vaccine supply chains, cold storage capacity, healthcare workforce distribution, and community attitudes toward vaccination. The environmental data used in our analysis, while comprehensive, represents long-term averages rather than real-time conditions during the vaccination campaign. We also lack direct measures of vaccine hesitancy or misinformation, which can significantly impede coverage in certain communities. Furthermore, while remote sensing data on nitrogen dioxide (NO_2_) levels and precipitation provided invaluable environmental insights, measurement imprecision may arise from spatial and temporal resolution gaps. K-Means clustering, for its part, hinges on predefined cluster counts, potentially oversimplifying subtler spatial patterns. These gaps point to the need for ongoing inquiry, particularly longitudinal studies that track how coverage responds to policy shifts, supply chain disruptions, or evolving public sentiment. Future research could also incorporate behavioral metrics, such as trust in health institutions, perceived vaccine efficacy, and the influence of social networks, to enrich our understanding of demand-side barriers.

## 5 CONCLUSIONS

Our study underscores the importance of recognizing that COVID-19 vaccination disparities in Kenya arise from structural injustices besides potential individual hesitancy or logistical oversights. Our spatially explicit methodology not only exposes the breadth of challenges, from sparse healthcare services in arid counties to financial hurdles that discourage vaccine uptake, but also offers a blueprint for data-driven policy interventions. By disentangling how factors such as bank access, sleeping room density, and seasonal floods intersect to shape coverage, we provide a roadmap for decision-makers seeking to break these cycles of disadvantage. This framework has resonance far beyond Kenya, as comparable inequities exist across diverse low- and middle-income countries grappling with vulnerable infrastructures, limited public spending, and heightened climate risks.

Embracing an integrated approach is crucial if global health agendas, including the Sustainable Development Goals and the African Union’s vision for vaccine self-reliance, are to be realized in communities traditionally left behind by mainstream development initiatives. Our findings highlight the urgency of aligning vaccination campaigns with broader objectives of universal health coverage, financial inclusion, and environmental resilience. Investing in mobile outreach, climate-resilient roads, and digital payment systems can help offset the cost of receiving a vaccine, thereby improving coverage in neglected regions. While such proposals demand political will and upfront expenditures, the long-term benefits, measured in reduced disease transmission, lower mortality, and avoided healthcare costs, are substantial. By adopting these practical steps, policymakers can create lasting improvements in equity and preparedness, ensuring that health interventions reach populations most at risk. Above all, this study urges a paradigm shift, effective pandemic responses hinge on tackling the social and geographic determinants of health, not just distributing vaccines. As the global community seeks to fortify its defenses against current and future pandemics, placing marginalized communities at the center of policy agendas stands as both a moral and a strategic imperative, essential for promoting resilient societies worldwide. Such an equity-focused philosophy can ultimately catalyze transformative progress in global health and sustainable development.

## Data Availability

Data are available in a public, open-access repository. The data that support the findings of this study are available from the Demographic and Health Surveys (http://www.measuredhs.com), but restrictions apply to the availability of these data, which were used under license for the current study and so are not publicly available. However, data are available from the authors on reasonable request and with the permission of Demographic and Health Surveys. We sought and were granted permission to use the core data set for this analysis by Measure DHS.

## DECLARATIONS

### Ethics approval

Procedures and questionnaires for standard Demographic and Health Surveys have been reviewed and approved by the ICF International Institutional Review Board (IRB). The ICF International IRB ensures that the survey complies with the US Department of Health and Human Services regulations for the protection of human subjects, while the host country IRB ensures that the survey complies with the laws and norms of the nation (http://dhsprogram.com/What-We-Do/Protecting-the-Privacy-of-DHS-Survey-Respondents.cfm). Institutional review board approval and informed consent were not necessary for this cross-sectional study because all data were deidentified and publicly available (Common Rule 45 CFR §46). This study follows the Strengthening the Reporting of Observational Studies in Epidemiology (STROBE) reporting guideline [**?** ].

## SUPPLEMENTARY FIGURES

**Figure S1.**
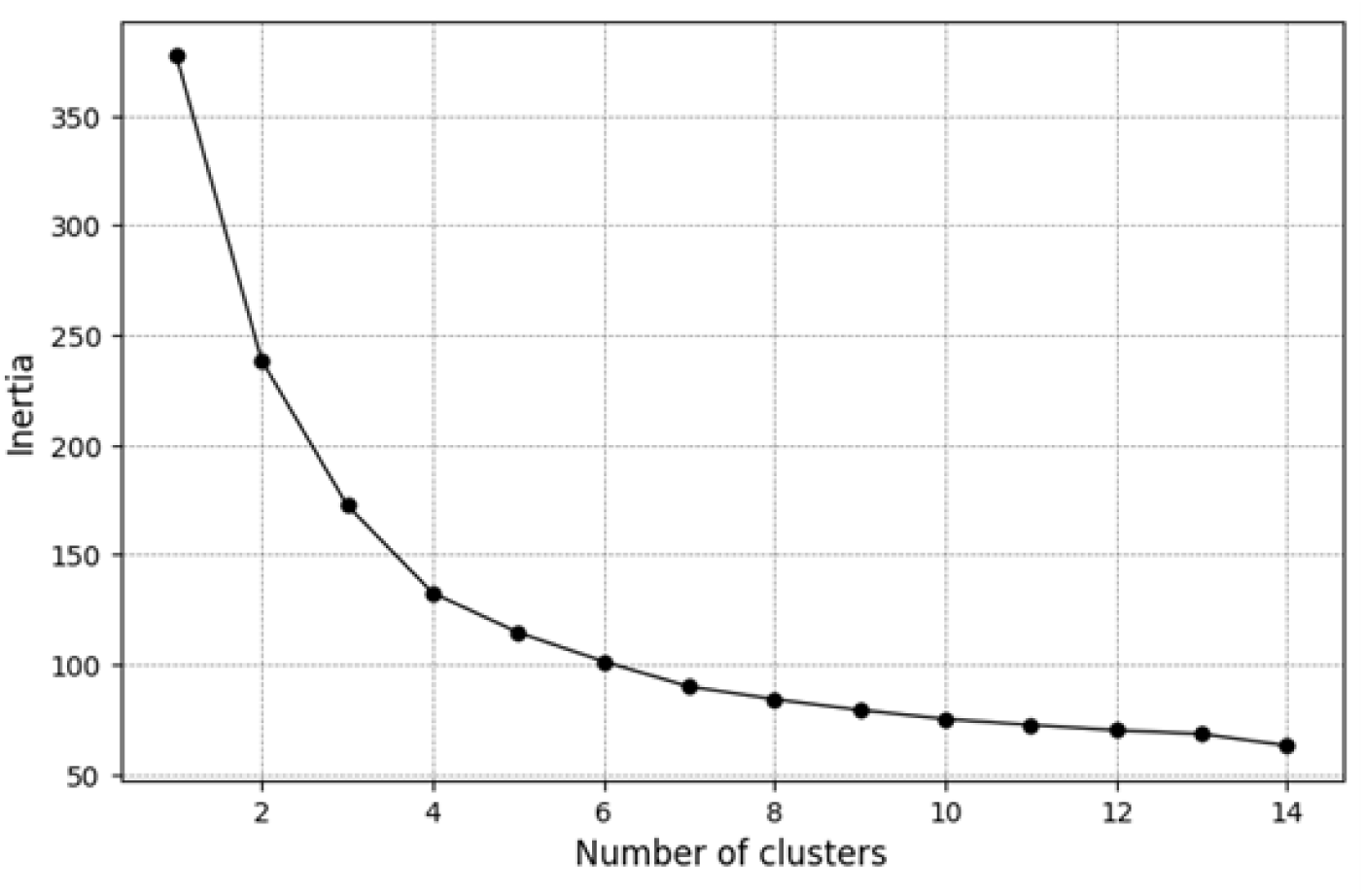
Elbow Test for Covariates of COVID-19 vaccination rates.

## SUPPLEMENTARY TABLES

**Table S1.** Socioeconomic variables included in the study, describing household-level characteristics and their measurement scales. These variables capture disparities in wealth, infrastructure, demographic structure, and access to basic services, all of which influence COVID-19 vaccination coverage.

| Variable Name | Description | Measurement | Justification |
| --- | --- | --- | --- |
| Wealth Index | Composite measure of household wealth based on asset ownership, housing materials, and access. | Ordinal (1: Poorest to 5: Richest) | Proxy for socio-economic status, associated with access to health services and vaccination uptake. |
| Bank Access | Proportion of households with access to formal banking services. | Binary (Yes/No) | Facilitates financial inclusion, enabling easier access to healthcare and vaccinations. |
| Type of Residence | Urban or rural classification of household location. | Binary (Urban/Rural) | Captures rural-urban disparities in access to healthcare infrastructure and vaccination centers. |
| Household Size | Number of members per household. | Numeric | Larger households may face logistical challenges in accessing vaccines, affecting coverage rates. |
| Age of Household Head | Age of the primary decision-maker in the household. | Numeric | Older household heads may influence vaccination attitudes and decision-making. |
| Child Ratio | Proportion of children under five relative to household size. | Numeric | Highlights demographic dependencies, with potential impacts on vaccination prioritization. |
| Roof Material | Classification of household roof material (e.g., natural, rudimentary, finished). | Ordinal (1: Natural to 3: Finished) | Proxy for household socio-economic status, influencing health-seeking behaviors. |
| Handwashing Place | Type of place used for handwashing in the household. | Ordinal (1: None to 3: Fixed Station) | Indicator of hygiene practices, critical for disease prevention and vaccine outreach campaigns. |
| Water Source | Type of drinking water source used by the household. | Ordinal (1: Surface to 4: Piped) | Reflects infrastructure and potential barriers to vaccine outreach, influencing health outcomes. |
| Social Assistance | Whether the household receives social assistance from the government or NGOs. | Binary (Yes/No) | Proxy for household reliance on external support, indicating vulnerability that may impact vaccination uptake. |
| Number of sleeping rooms per household | Number of sleeping rooms in the household. | Numeric | Indicator of crowding, which can exacerbate disease spread and influence vaccination priorities. |
| Gender Ratio | Gender of the primary decision-maker in the household. | Binary (Man/Woman) | Gender household heads may influence vaccination attitudes and decision-making. |
| Toilet Type | Type of toilet used in the household. | Ordinal (1: None to 3: improved toilet) | Indicator of hygiene practices, critical for disease prevention. |
| Travel Time by Vehicle | Travel time to the nearest health facility using a motor vehicle. | Numeric | Proxy for level access to health services. |
| Travel Time Without Vehicle | Travel time to the nearest health facility without using a motor vehicle. | Numeric | Proxy for level access to health services. |

**Table S2.** Health-related variables included in the study, detailing individual and community-level health metrics. These variables reflect the burden of infectious diseases, preventive practices, and healthcare access, which are critical for understanding vaccination patterns.

| Variable Name | Description | Measurement | Justification |
| --- | --- | --- | --- |
| HIV Prevalence | Proportion of the population living with HIV in the area. | Numeric | Captures the burden of chronic illness, which may influence vaccine prioritization and delivery. |
| Malaria Prevalence | Proportion of the population affected by malaria in the area. | Numeric | Reflects regional health challenges that may interact with vaccination priorities and healthcare access. |
| Child Immunization Rate | Proportion of children in the household fully vaccinated for standard childhood diseases. | Numeric | Proxy for healthcare access and attitudes toward vaccination, predicting COVID-19 vaccine acceptance. |
| Bednet Use | Proportion of households using insecticide-treated bednets. | Numeric | Indicator of malaria prevention efforts, reflecting access to public health interventions. |
| Indoor Smoking | Frequency of smoking inside the household. | Ordinal (0: Never to 4: Daily) | Assesses respiratory health risks, which may affect vaccine prioritization for COVID-19. |
| Immunization Rate | Community-level rate of routine vaccinations among children. | Numeric | Proxy for healthcare delivery effectiveness and general acceptance of vaccines. |
| COVID-19 Incidence | Number of people tested positive for COVID-19. | Numeric | Proxy for healthcare access, predicting COVID-19 vaccine acceptance. |
| COVID-19 Deaths | Number of people died from COVID-19. | Numeric | Reflects regional health challenges that predicting COVID-19 vaccine acceptance. |
| Cooking Fuel | Type of cooking fuel. | Ordinal (1: Biomass to 5: electricity) | Assesses respiratory health risks, which may affect vaccine prioritization for COVID-19. |

**Table S3.**
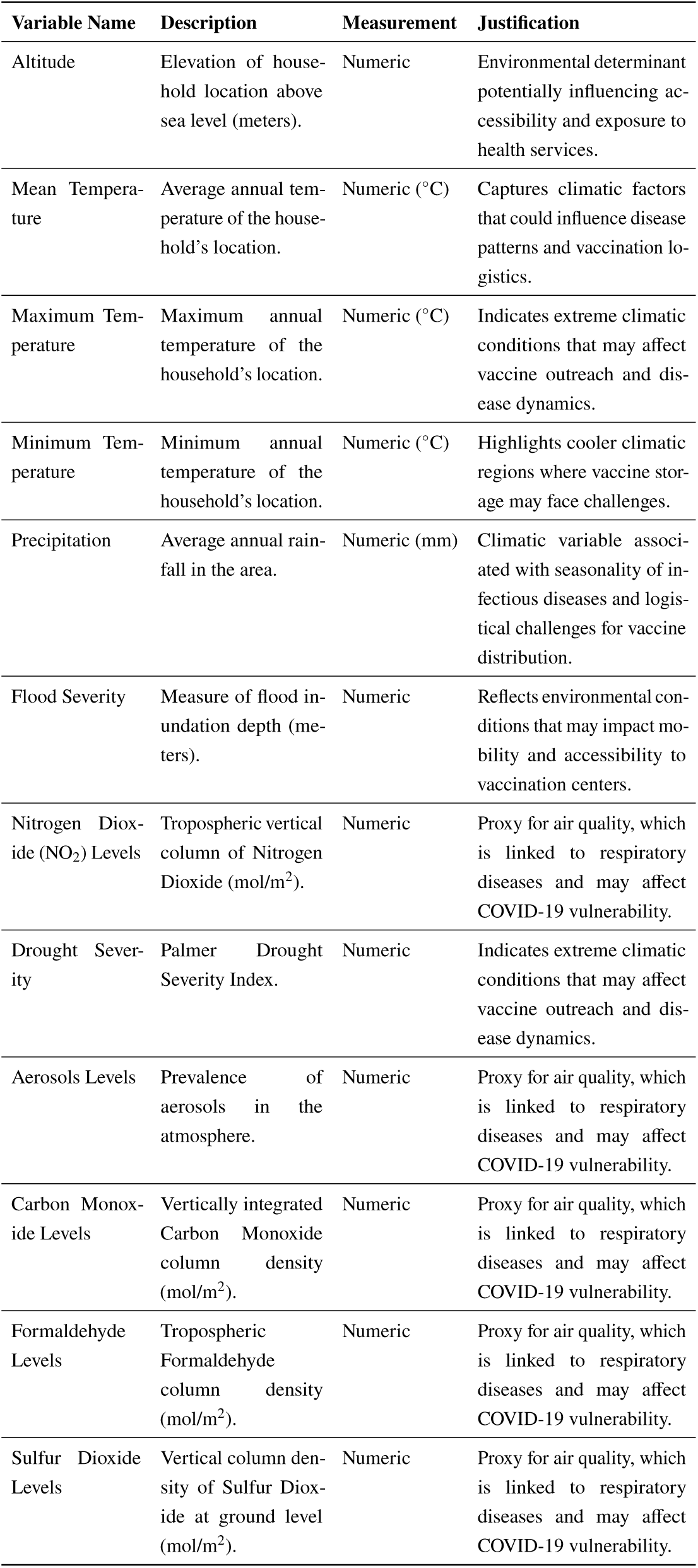
Environmental variables included in the study, describing climatic and geographic characteristics of household locations. These factors influence disease dynamics, logistical challenges, and accessibility to health services, shaping vaccination uptake.

| Variable Name | Description | Measurement | Justification |
| --- | --- | --- | --- |
| Altitude | Elevation of household location above sea level (meters). | Numeric | Environmental determinant potentially influencing accessibility and exposure to health services. |
| Mean Temperature | Average annual temperature of the household's location. | Numeric (°C) | Captures climatic factors that could influence disease patterns and vaccination logistics. |
| Maximum Temperature | Maximum annual temperature of the household's location. | Numeric (°C) | Indicates extreme climatic conditions that may affect vaccine outreach and disease dynamics. |
| Minimum Temperature | Minimum annual temperature of the household's location. | Numeric (°C) | Highlights cooler climatic regions where vaccine storage may face challenges. |
| Precipitation | Average annual rainfall in the area. | Numeric (mm) | Climatic variable associated with seasonality of infectious diseases and logistical challenges for vaccine distribution. |
| Flood Severity | Measure of flood inundation depth (meters). | Numeric | Reflects environmental conditions that may impact mobility and accessibility to vaccination centers. |
| Nitrogen Dioxide (NO<sub>2</sub>) Levels | Tropospheric vertical column of Nitrogen Dioxide (mol/m<sup>2</sup>). | Numeric | Proxy for air quality, which is linked to respiratory diseases and may affect COVID-19 vulnerability. |
| Drought Severity | Palmer Drought Severity Index. | Numeric | Indicates extreme climatic conditions that may affect vaccine outreach and disease dynamics. |
| Aerosols Levels | Prevalence of aerosols in the atmosphere. | Numeric | Proxy for air quality, which is linked to respiratory diseases and may affect COVID-19 vulnerability. |
| Carbon Monoxide Levels | Vertically integrated Carbon Monoxide column density (mol/m<sup>2</sup>). | Numeric | Proxy for air quality, which is linked to respiratory diseases and may affect COVID-19 vulnerability. |
| Formaldehyde Levels | Tropospheric Formaldehyde column density (mol/m<sup>2</sup>). | Numeric | Proxy for air quality, which is linked to respiratory diseases and may affect COVID-19 vulnerability. |
| Sulfur Dioxide Levels | Vertical column density of Sulfur Dioxide at ground level (mol/m<sup>2</sup>). | Numeric | Proxy for air quality, which is linked to respiratory diseases and may affect COVID-19 vulnerability. |

